# Healthy lifestyle and prostate cancer risk in the Million Veteran Program

**DOI:** 10.1101/2022.07.09.22277437

**Authors:** Meghana S. Pagadala, Asona Lui, Julie Lynch, Roshan Karunamuni, Kyung Min Lee, Anna Plym, Brent S. Rose, Hannah Carter, Adam S. Kibel, Scott L. DuVall, J. Michael Gaziano, Matthew S. Panizzon, Richard L. Hauger, Tyler M. Seibert

## Abstract

**Importance:** Prostate cancer (PCa) risk is understood to be mostly unmodifiable and inherited, but there is evidence that environmental and behavioral factors may also contribute. A recent study of health professional cohorts suggests a healthy lifestyle can mitigate a high inherited risk of lethal PCa. It is unknown how modifiable factors affect PCa risk in more diverse populations.

**Objective:** To determine the effects of healthy lifestyle on PCa risk when accounting for race/ethnicity, family history, and genetic risk in a diverse population.

**Design:** Retrospective analysis of the Million Veteran Program (MVP), a cohort study conducted 2011-2021.

**Setting:** National, population-based cohort of United States military veterans.

**Participants:** All veterans were eligible to participate. 590,750 male participants were available for this study; 281,923 completed the lifestyle survey.

**Exposures:** Smoking status, exercise (strenuous activity ≥ 2 days per week), family history, race, and ethnicity (all self-reported at enrollment). PSA testing and body mass index (BMI, obtained from clinical records). Genetic risk (assessed via a polygenic hazard score using genotype data).

**Main Outcomes:** Age at diagnosis of PCa, diagnosis of metastatic PCa, and death from PCa were assessed via Cox proportional hazards models.

**Results:** On univariable analysis, exercise and not smoking were associated with reduced prostate cancer risk (PCa incidence, metastasis, and death), while higher BMI was associated only with reduced PCa death. Not smoking was independently associated with reduced metastatic PCa (HR 0.54, 95% CI 0.49–0.59, p<10^−16^) and fatal PCa (HR 0.35, 95% CI 0.30– 0.41, p<10^−16^) when accounting for exercise, BMI, family history, genetic risk, and race/ethnicity. Exercise was not independently associated with any PCa endpoints. Higher BMI was independently associated with reduced fatal PCa (HR 0.97 95% CI 0.96–0.99, p<0.01). Lifetime cause-specific mortality at age 85 was lower for non-smokers (vs. smokers) among Non-Hispanic White men (0.7% vs. 1.8%) and among Black men (1.7% vs. 6.1%).

**Conclusions:** Smoking is independently associated with increased risk of metastatic and fatal PCa among men of diverse race and ethnicity. This demonstrates that lifestyle factors can mitigate the risk of PCa death in high-risk men.

**Key points:** *Question:* Is a healthy lifestyle associated with reduced risk of fatal prostate cancer in a diverse population?

*Findings:* In a population-based cohort study of 281,923 US military veterans, non-smokers had a slight reduction in risk of being diagnosed with prostate cancer and 0.35 times risk of dying from it, a significant association. Lifetime cause-specific mortality at age 85 was lower for non-smokers among Non-Hispanic White men (0.7% vs. 1.8%) and among Black men (1.7% vs. 6.1%).

*Meaning:* Men who smoke are more likely to develop fatal prostate cancer.

## Introduction

Prostate cancer (PCa) risk is understood to be mostly inherited and not modifiable. The strongest known predictors of risk are age, race/ethnicity (especially Black race), family history, and specific genetic elements.^1-7^ However, there is growing evidence of modifiable PCa risk factors, including environmental elements and behavior.^8^ Meta-analyses of observational studies have identified an association between tobacco use, low physical activity, and obesity with increased risk of PCa progression and PCa-related death.^8-11^ Specific dietary factors have also been linked to increased risk of PCa or PCa progression.^12,13^

An intriguing recent study of two cohorts of health professionals suggested a healthy lifestyle can mitigate a high inherited risk of lethal PCa.^14^ Healthy lifestyle, including regular exercise and a wholesome diet (low in saturated fats but high in fruits, vegetables, low-fat dairy and fish) is also known to reduce the risks of cardiovascular disease, colorectal cancer, and breast cancer.^15-17^ However, it is not known whether the protective effect of a healthy lifestyle on PCa risk holds true for more diverse populations, especially when accounting for race/ethnicity and family history.

We investigated the association of modifiable lifestyle and environmental factors on PCa risk in a diverse population when accounting for common major inherited risk factors (family history and genetic risk) and for race/ethnicity. We used data from the Department of Veteran Affairs (VA) Million Veteran Program (MVP), a population-based cohort with genotyping, long-term follow-up, and linked clinical records for over 870,000 participating US veterans. The MVP is one of the largest and most diverse electronic health record-linked biobanks in the world, with a unique structure that allows for detailed investigation into the interactions between inherited risk and lifestyle.^18^ We tested previously reported indicators of a healthy lifestyle (not smoking, healthy weight, and exercise) for associations with age at PCa diagnosis, age at diagnosis of PCa metastasis, and lifetime PCa-specific mortality. We hypothesized that a healthy lifestyle is associated with reduced risk of metastatic and fatal prostate cancer.

## Methods

### Participants

We obtained data from MVP for individuals recruited from 63 Veterans Affairs Medical Centers across the United States (US) beginning in 2011. All veterans were eligible for participation in MVP. Study participation included consenting to access the participant’s electronic health records for research purposes. The MVP received ethical and study protocol approval from the VA Central Institutional Review Board in accordance with the principles outlined in the Declaration of Helsinki. Only men were included in this PCa study. Participant demographics are shown in **Table 1**.

**Table 1:**
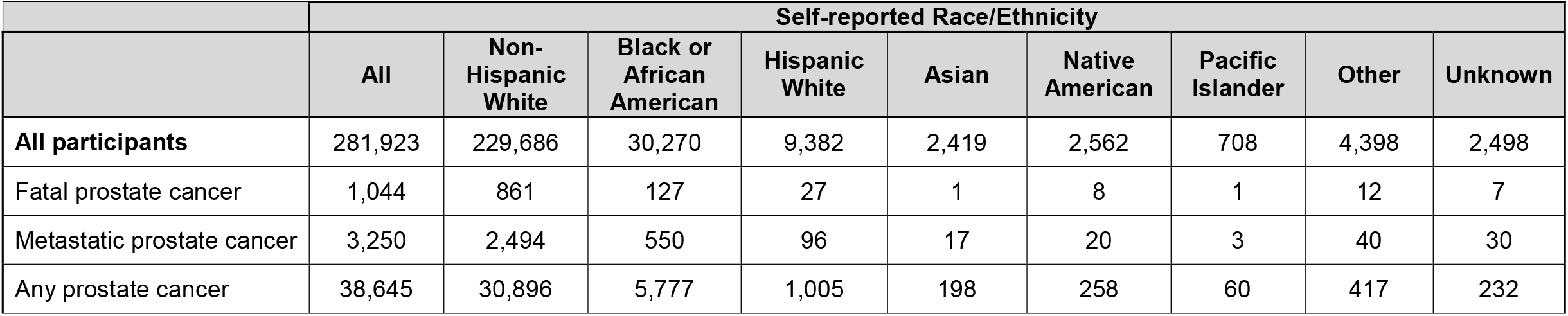
Participant characteristics for self-reported race/ethnicity groups. Numbers indicate the number of participants in each group available for lifestyle analysis.

### Healthy lifestyle factors

281,923 of the 590,750 MVP participants completed a questionnaire with nearly 400 detailed behavioral and diet questions. We included lifestyle factors similar to those reported in previous studies^14,15^: smoking, body mass index (BMI), and exercise (strenuous physical activity ≥ 2 days per week). BMI was calculated from the participants’ baseline height and weight obtained at time of enrollment in MVP. Analysis was restricted to the 281,923 participants that responded to the relevant lifestyle questions, resulting in no missing data for smoking status or exercise. All participants had height and weight information available for BMI calculation.

### Clinical data extraction

Comprehensive description of clinical endpoints used for PCa Cox proportional hazards analysis were described previously.^19^ Briefly, PCa diagnosis, age at diagnosis, PSA tests, and date of last follow-up were retrieved from the VA Corporate Data Warehouse based on ICD codes and VA Central Cancer Registry data. Age at diagnosis of metastatic PCa indicated the age of the participant when diagnosed with either nodal or distant metastases as determined through a validated natural language processing tool.^27^ Fatal PCa information was determined from National Death Index. Participants with ICD10 code “C61” as underlying cause of death were considered to have died from PCa. Family history was recorded as either the presence or absence of (one or more) first-degree relatives with PCa. Among the 281,923 participants eligible for analysis, less than 1% had never received a PSA test in the VA system.

### Genetic risk: polygenic hazard score (PHS290)

Blood sampling and DNA extraction were conducted by MVP as described previously.^1,20^ The MVP 1.0 genotyping array contains a total of 723,305 variants, enriched for low frequency variants in African and Hispanic populations and variants associated with diseases common to the VA population.^25^ Details on quality control and imputation have been described previously.^20^

To assess genetic risk, we calculated a previously developed and validated polygenic hazard score using 290 common genetic variants (PHS290) that reliably stratifies men for age-dependent genetic risk of PCa and is associated with PCa, metastatic PCa, and PCa death.^5^ Details of PHS290 calculation in MVP are described elsewhere. ^1,5,19^ PHS290 performs well in diverse datasets and is independently associated with PCa risk.^1,5,19^

### Cox proportional hazards analysis

We used Cox proportional hazards models to evaluate the association of healthy lifestyle factors with three clinical endpoints: age at diagnosis of PCa, age at diagnosis of metastatic PCa, and age at death from PCa. We also analyzed self-reported racial/ethnic subgroups. As very few participants identified as being of both Black race and Hispanic ethnicity, we included these in a single category for Black or African-American race (Black). Where individuals did not meet the endpoint of interest, we censored at age at last follow-up. We also tested each lifestyle factor on univariable analysis.

To assess for independent association of lifestyle with PCa endpoints when accounting for inherited risk, we used multivariable Cox proportional hazards models. We have previously shown that combining race/ethnicity, family history, and PHS290 provides better risk stratification than any single variable alone.^1^ Here, we tested lifestyle factors for associations in models with race/ethnicity and family history, without and with PHS290. For race/ethnicity hazard ratios, we used Non-Hispanic White as reference. For PHS290, we illustrated the effect size via the hazard ratio for the highest 20% vs. lowest 20% of genetic risk (HR80/20) and between other strata of PHS290. These percentiles refer to previously defined thresholds of PHS290 derived from European men unaffected by PCa and <70 years old. Further details and assessment of alternate strategies in the MVP population is described elsewhere.^5,19^ We assessed statistical significance with two-tailed alpha at 0.01.

### PSA testing

Screening has been shown in a large, randomized trial to increase PCa incidence and reduce cause-specific mortality^21^, raising the possibility that PSA testing may confound any impact of healthy lifestyle. The reason for PSA tests in MVP participants is unknown, but we were able to count the number of PSA tests each participant underwent. We tested for associations between lifestyle factors and number of PSA tests for a given participant by performing multivariable linear regression and using race/ethnicity, family history, and each of the lifestyle factors as predictive variables. This analysis was performed in men who did not have prostate cancer to avoid confounding by diagnostic PSA tests.

### Dietary factors: exploratory analysis

While prior studies have also reported associations for specific dietary habits, it was not immediately clear how to best incorporate the dietary information available in MVP. We therefore conducted an exploratory analysis using responses to 126 dietary questions (at MVP enrollment), testing for univariable association with the three PCa endpoints. Cox model *p*-values and hazard ratios (with 95% confidence intervals) were tabulated to inform future studies.

## Results

### Participant characteristics

Median age at MVP enrollment was 68 years (interquartile range 62-74). Median age at last follow-up was 72 (66-78). The distribution of lifestyle factors across racial/ethnic groups is shown in **Supplementary Figure 1**.

### Association of lifestyle factors with PCa endpoints

On univariable analysis of lifestyle factors (**Supplementary Table 1**), non-smokers had lower risk of PCa (HR 0.89, [0.86–0.92], *p*<10^−11^), metastatic PCa (HR 0.50, [0.46–0.55], *p*<10^−16^), and fatal PCa (HR 0.33, [0.28–0.39], *p*<10^−16^). Exercise was associated with lower risk of metastatic PCa (HR 0.74, [0.68–0.81], p<10^−10^) and fatal PCa (HR 0.74, [0.64–0.86], p<10^−3^), as well as slightly lower risk of any PCa (HR 0.95, [0.93–0.98], p<10^−3^). Higher BMI was not associated with any PCa (*p*=0.052) or metastatic PCa (*p*=0.13) but was associated with lower risk of fatal PCa (*p*<10^−6^). Cause-specific cumulative incidence curves for metastatic PCa and PCa death diverged for smokers and non-smokers (**Figure 1**).

**Figure 1.**
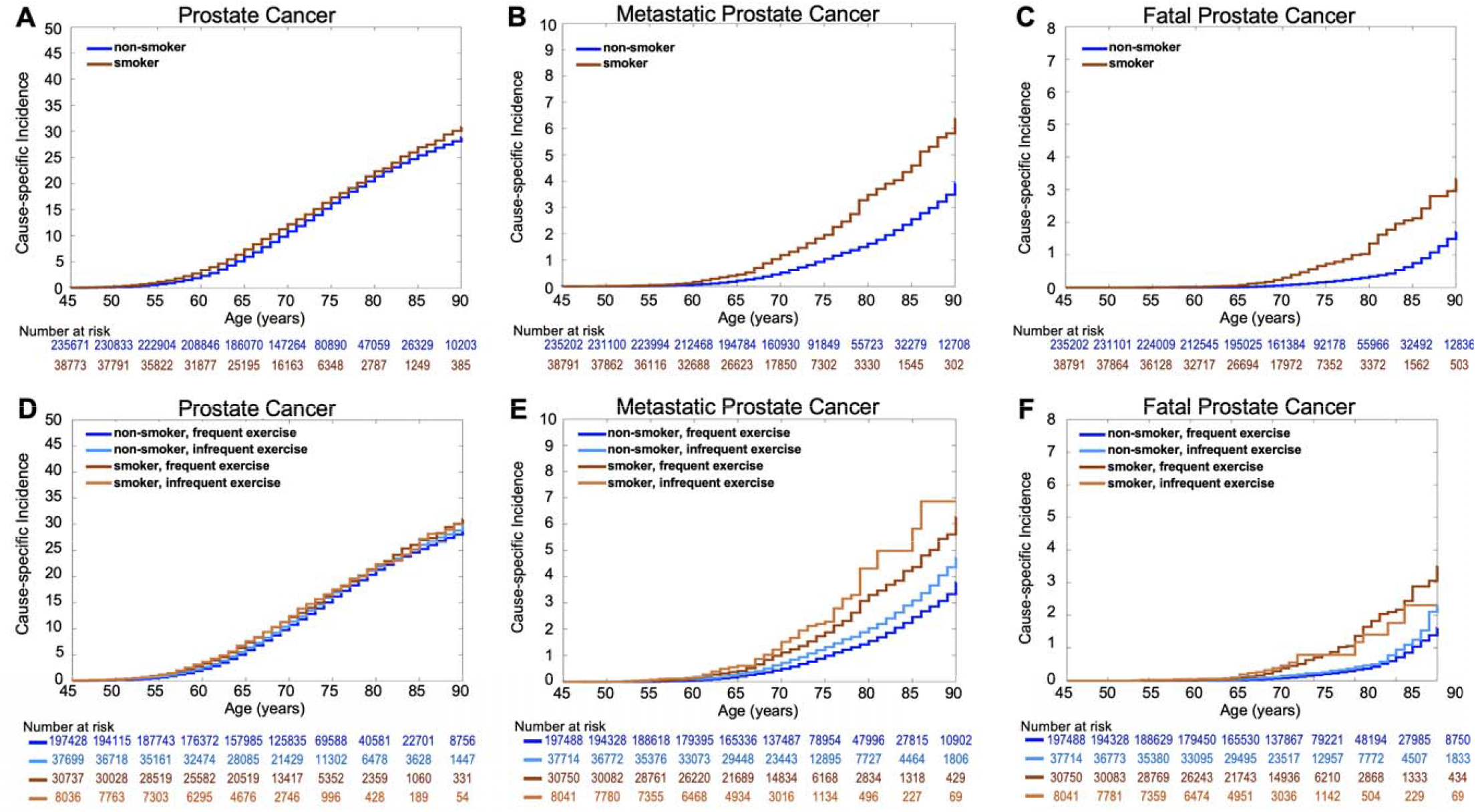
Million Veteran Program (MVP) Cause-specific Cumulative Incidence based on lifestyle factors. Cause-specific cumulative incidence within MVP, stratified by smoking status (top row) and by both smoking status and exercise (bottom row) for **(A, D)** all prostate cancer, **(B, E)** metastatic prostate cancer, and **(C, F)** fatal cancer.

Black participants who did not smoke had no reduction in PCa incidence (HR 0.98, [0.92–1.05], *p*=0.60) but had reduced risk of metastatic PCa (HR 0.62, [0.51–0.77], *p*<10^−5^) and fatal PCa (HR 0.31, [0.21–0.48], *p*<10^−8^) (**Supplementary Table 1**). Absolute risk reduction was greater among Black men (**Figure 2**). Lifetime cause-specific mortality at age 85 was lower for non-smokers among Non-Hispanic White men (0.7% vs. 1.8%) and among Black men (1.7% vs.6.1 %). Metastatic PCa rates at age 85 were also lower for non-smokers among Non-Hispanic White men (2.3% vs. 4.0%) and among Black men (6.4% vs. 9.2%).

**Figure 2.**
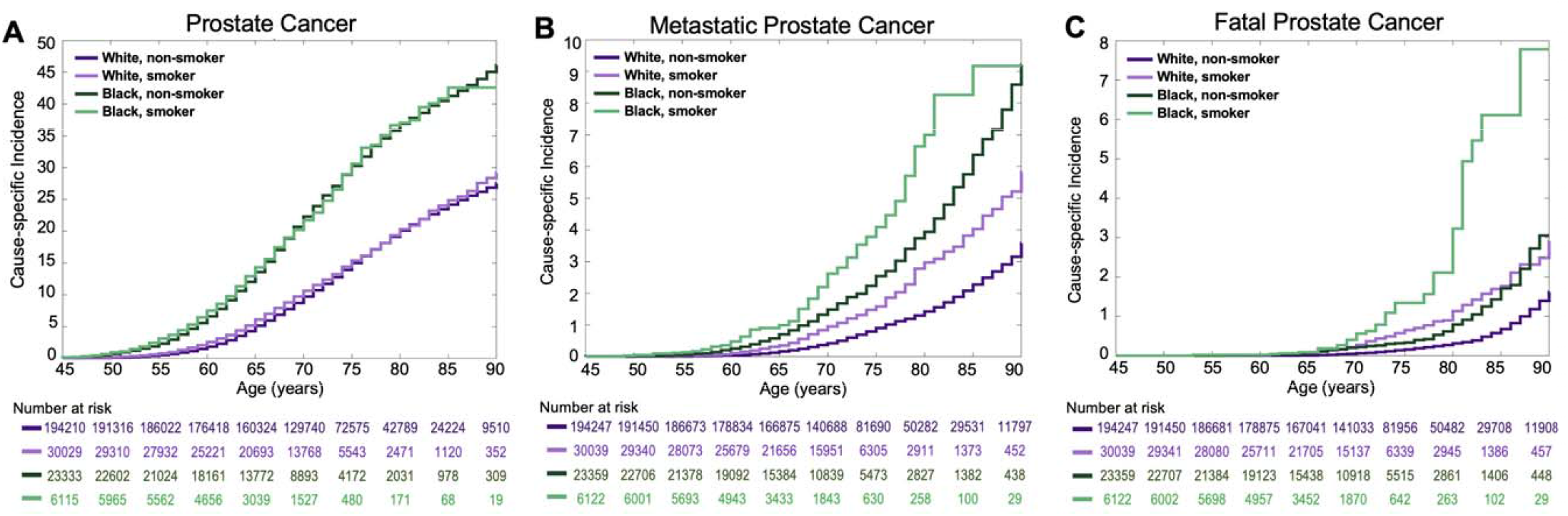
Million Veteran Program (MVP) Cause-specific Cumulative Incidence based on smoking status for White and Black participants. Cause-specific cumulative incidence within MVP, stratified by smoking status and also by self-reported race for **(A)** all prostate cancer, **(B)** metastatic prostate cancer, and **(C)** fatal cancer. “White” indicates Non-Hispanic White participants, and “Black” indicates Black and Hispanic Black participants.

When accounting for race/ethnicity and family history in multivariable analysis, not smoking was independently associated with reduced PCa (HR 0.94, [0.91–0.98], *p*<0.001), metastatic PCa (HR 0.54, [0.49–0.59], *p*<10^−16^) and fatal PCa (HR 0.35, [0.3–0.41], *p*<10^−16^) (**Table 2**). Higher BMI was associated with reduced fatal PCa risk (HR 0.97 95% CI 0.96–0.99, p<0.01) but not with risk of PCa diagnosis or metastatic PCa. When excluding both underweight (BMI <18.5) and very obese participants (BMI >35)^22,23^, higher BMI was associated with increased risk of any PCa, but the association with lower risk of fatal disease remained (**Supplementary Table 2**). Exercise was not independently associated with PCa risk on multivariable analysis. Accounting for genetic risk (PHS290) as well as race/ethnicity and family history had minimal impact on the strong associations with smoking status (**Table 3**). However, absolute risk reduction was greater among men with high genetic risk (PHS290 >80^th^ percentile, as defined previously^5^) (**Supplementary Figure 2**). Among men with high genetic risk, lifetime cause-specific mortality at age 85 was 1.4% for non-smokers vs. 4.3% for smokers (compared to 0.7% vs. 2.1% among the whole population). Likewise, among men with high genetic risk, metastatic PCa rates were 4.4% for non-smokers vs. 8.9% for smokers (compared to 2.6% vs. 4.6% among the whole population).

**Table 2:** Multivariable models combining self-reported Race/Ethnicity, Family History (FH), and lifestyle factors for three prostate cancer clinical endpoints. Cox proportional hazards results for association with age at death from prostate cancer, at diagnosis of metastatic prostate cancer and age at diagnosis with prostate cancer. *P*-values reported are from multivariable models using self-reported race/ethnicity, family history and three lifestyle factors (non-smoker, BMI, and exercise). Hazard ratios for race/ethnicity were estimated using Non-Hispanic White as reference. Hazard ratios for family history were for one or more first-degree relatives diagnosed with prostate cancer. Numbers in brackets are 95% confidence intervals. Significant predictors in the multivariable model are indicated by *(*p*<0.01), **(p<10^−6^) and *** (p<10^−16^).

| Clinical Endpoint | Race/Ethnicity |  |  |  |  |  |  | FH | Lifestyle |  |  |
| --- | --- | --- | --- | --- | --- | --- | --- | --- | --- | --- | --- |
|  | Black | Hispanic | Asian | Native American | Pacific Islander | Other | Unknown |  | Non-Smoker | BMI | Exercise |
| Fatal Prostate Cancer | 2.35<br>[1.89-2.85]*** | 1.13<br>[0.68-1.63] | 0.21<br>[0.0-0.7] | 1.25<br>[0.26-2.45] | 1.4<br>[0.0-4.63] | 1.77<br>[0.79-2.9] | 0.0<br>[0.0-0.0] | 2.02<br>[1.63-2.46]*** | 0.35<br>[0.3-0.41]*** | 0.97<br>[0.96-0.99]* | 0.85<br>[0.7-1.06] |
| Metastatic Prostate Cancer | 2.75<br>[2.48-3.04]*** | 1.25<br>[0.96-1.55] | 0.89<br>[0.45-1.41] | 1.11<br>[0.65-1.67] | 0.8<br>[0.0-2.02] | 1.67<br>[1.16-2.23]* | 1.71<br>[0.7-2.87] | 1.74<br>[1.55-1.97]*** | 0.54<br>[0.49-0.59]*** | 1.0<br>[0.99-1.01] | 0.9<br>[0.8-1.03] |
| Prostate Cancer | 2.3<br>[2.23-2.38]*** | 1.07<br>[0.99-1.15] | 0.9<br>[0.78-1.04] | 1.08<br>[0.94-1.21] | 1.07<br>[0.75-1.41] | 1.12<br>[1.01-1.23] | 0.89<br>[0.68-1.12] | 1.93<br>[1.87-1.98]*** | 0.94<br>[0.91-0.98]* | 1.0<br>[1.0-1.0] | 1.0<br>[0.97-1.04] |

**Table 3:**
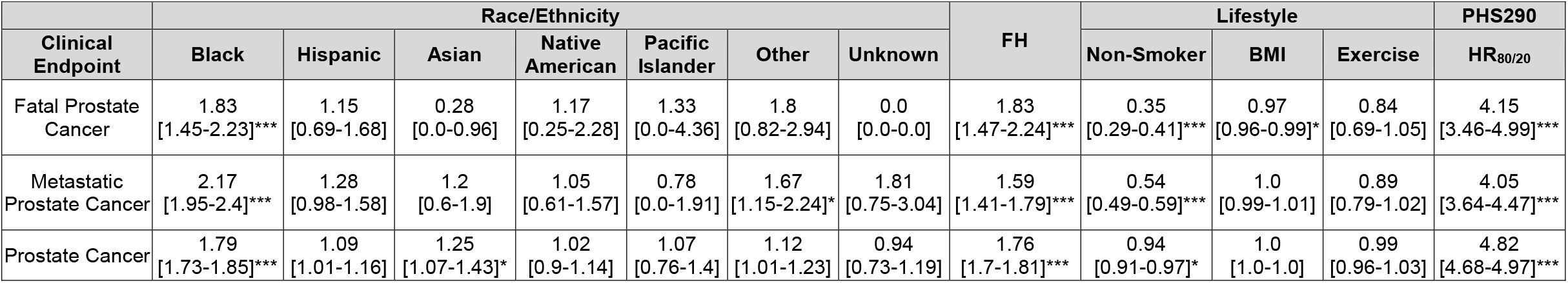
Multivariable models combining self-reported Race/Ethnicity, Family History (FH), Lifestyle factors, and PHS290 for three prostate cancer clinical endpoints. Cox proportional hazards results for association with age at death from prostate cancer, at diagnosis of metastatic prostate cancer and age at diagnosis with prostate cancer. For PHS290, effect size was illustrated via the hazard ratio (HR_80/20_) for the highest 20% vs. lowest 20% of genetic risk. Hazard ratios for race/ethnicity were estimated using Non-Hispanic White as the reference. Hazard ratios for family history were for one or more first-degree relatives diagnosed with prostate cancer. Numbers in brackets are 95% confidence intervals. Significant predictors in the multivariable model are indicated by *(*p*<0.01), **(p<10^−6^) and *** (p<10^−16^).

### PSA testing

Most MVP participants (>99%) underwent PSA testing, with many undergoing multiple PSA tests (**Supplementary Figure 3)**. Several variables were independently associated with the number of PSA tests reported per MVP participant, including participant race (**Supplementary Table 3**). Not smoking and frequent exercise were independently associated with slightly fewer PSA tests, while higher BMI was associated with slightly more PSA tests.

### Dietary factors: exploratory analysis

In the exploratory analysis of 126 baseline survey questions, 84, 73 and 41 variables had univariable statistically significant association with PCa diagnosis, metastatic PCa, and PCa death, respectively (**Supplementary Table 4**). Highly significant variables associated with increased PCa risks included vitamin D and calcium supplementation; consumption of fried foods; processed meats; sugary drinks/snacks; and daily alcohol use.

## Discussion

In a diverse, population-based cohort, healthy lifestyle mitigated inherited risk of PCa—and especially risk of metastatic and fatal PCa. Not smoking and strenuous physical activity were each associated with reductions in any PCa, metastatic PCa, and fatal PCa on univariable analysis, while lower BMI was associated with increased fatal PCa only. Smoking showed a meaningful independent association with reduced metastatic and fatal PCa (and a slight reduction in PCa incidence) when accounting for strong associations with family history, race/ethnicity, and genetic risk. On multivariable analysis, smoking approximately doubled the lifetime risk of metastatic PCa and tripled the lifetime risk of fatal PCa.

These findings build on those of a previous study that found healthy lifestyle was associated with reduced lethal PCa (combining metastatic and fatal PCa) among White health professionals with high genetic risk.^14^ The present work demonstrates healthy lifestyle mitigates inherited risk across the full spectrum of PHS290 and across diverse racial/ethnic groups, including Black men. This is important because Black men are, on average, at high risk of PCa mortality. We observed a larger absolute difference in fatal PCa between smokers and non-smokers among Black men.

We note that relationships between ancestry, family history, and genetic risk are especially complicated in the US due to a variety of factors, including but not limited to high diversity, structural racism, social biases, and social determinants of health. To mirror current clinical guidelines, and because genetic ancestry is not available during most clinical encounters, we used self-reported ancestry for the primary analyses. Self-reported race/ethnicity, as a social construct, may also more faithfully reflect the social determinants of health associated with PCa risks and outcomes.^24-27^ Previously, we found that using genetic ancestry or the top 10 principal components of genetic variation rather than self-reported ancestry yielded essentially unchanged results.^1,5^ Due to small numbers, we considered Hispanic ethnicity separately within White men but not within other races. Recent work suggests significant differences in PCa risk among US Hispanic men based on country of origin.^7,28^ Future work would benefit from the ability to conduct subgroup comparisons among Hispanic/Latino US men. As with race/ethnicity, family history of PCa is also likely measuring a combination of genetic risk, social determinants of health, and even learned behaviors like smoking and exercise habits.

The oft-unknown impact of PSA testing is a limitation of many epidemiological studies of PCa mortality, including the prior study of lifestyle^14^. If, for example, non-smokers were screened more often, we would potentially expect higher PCa incidence and lower PCa mortality in that group. However, we did not find increased PCa incidence among non-smokers (incidence was slightly lower). Moreover, on multivariable analysis, we found non-smokers had significantly *less* PSA tests than smokers. Both findings mitigate concerns that screening differences could explain the increased PCa mortality among smokers.

Higher BMI was independently associated with reduced risk of fatal PCa. When limiting analysis to only participants with BMI 18.5-35, higher BMI was independently associated with increased PCa diagnosis but the association between lower BMI and fatal PCa remained. An association between high BMI and/or higher percent of body fat and PCa risk has been controversial in prior studies, with reports of both positive and negative associations with fatal prostate PCa^29-31^. Further work will be necessary to understand the complex relationship between a healthy weight and PCa risk.

Because the MVP questionnaire did not include the same questions used in similar PCa studies^14^ and a validated diet score could not be reproduced with the available data, we did not include diet in the multivariable analysis. On exploratory univariable analysis, the association of individual diet components with PCa risks were generally consistent with prior studies in other datasets.^8,12,13^ Consumption of processed meat products, eggs, fried foods, whole milk and sugary beverages were associated with increased PCa risks. Conversely, consumption of nuts, coffee, chocolate, and tomatoes were associated with lower PCa risks. Supplementation with multi-vitamins and Omega 3 fatty acids were associated with lower risk of PCa metastasis and cause-specific death, while supplementation with Calcium and Vitamin D were associated with increased risks. Notably, this analysis assumed a linearity of the raw data. Future studies should explore how different thresholds of each of these variables impact PCa diagnosis and clinical outcomes.

This study was conducted using the MVP dataset and so the results and conclusions may not be generalizable beyond the VA population. As sequencing for rare pathogenic mutations was not performed, it was not possible to assess the impact of a healthy lifestyle on risk arising from, for example, germline *BRCA2* mutations. It is possible some participants underwent PSA testing outside the VA, which we could not capture here. Finally, lifestyle factors were only measured at the time of MVP enrollment, and so does not capture fluctuations in behaviors and lifestyle over time—e.g., changes in exercise habits. Future work should evaluate how longitudinal lifestyle factors (duration, age of adoption, timing before or after diagnosis, years since quitting smoking etc.) impact a man’s PCa risk.

## Conclusions

Not smoking and strenuous physical activity are associated with reduced metastatic and fatal PCa. Smoking is independently associated with increased risk of metastatic and fatal PCa when accounting for major non-modifiable factors (family history, Black race, and high genetic risk). Prospective studies should investigate whether healthy lifestyle interventions can reduce PCa risks, especially in Black men and men with high inherited risk.

## Data Availability

All data produced in the present study are available upon reasonable request to the authors. Access to the raw data in the Million Veterans Program (MVP) is available upon special request through the US Department of Veteran Affairs.

## Acknowledgements

This research used data from the Million Veteran Program, Office of Research and Development, Veterans Health Administration. This research was supported by the Million Veteran Program MVP022 award # I01 CX001727 (PI: Richard L. Hauger MD). This publication does not represent the views of the Department of Veterans Affairs or the United States Government. Dr. Hauger was additionally funded by the VISN-22 VA Center of Excellence for Stress and Mental Health (CESAMH) and National Institute of Aging RO1 grant AG050595 (*The VETSA Longitudinal Twin Study of Cognition and Aging VETSA 4)*. This research was supported by VA MVP022. Meghana S. Pagadala was supported by the National Institutes of Health (#1F30CA247168, #T32CA067754). Asona J. Lui was supported by the Grillo-Marxuach Family Fellowship at the Moores Cancer Institute of UC San Diego. Anna Plym was supported by the Prostate Cancer Foundation (Young Investigator Award) and the Swedish Cancer Society (Fellowship). Tyler Seibert and Roshan Karunamuni were supported by the National Institutes of Health (NIH/NIBIB #K08EB026503), the Prostate Cancer Foundation, and the University of California (#C21CR2060).

## Data Availability Statement

It is not possible for the authors to directly share the individual-level data that were obtained from the MVP due to constraints stipulated in the informed consent. Anyone wishing to gain access to this data should inquire directly to MVP at. The data generated from our analyses are included in the manuscript main text, tables, and figures and online supplementary materials. The code used for analyses is available at https://github.com/precimed/MVP-PCa-PHS.

## References

1. Pagadala MS, Lynch J, Karunamuni R, et al. Polygenic risk of any, metastatic, and fatal prostate cancer in the Million Veteran Program. medRxiv. 2022:2021.09.24.21264093. doi:10.1101/2021.09.24.21264093

2. Seibert TM, Fan CC, Wang Y, et al. Polygenic hazard score to guide screening for aggressive prostate cancer: development and validation in large scale cohorts. (1756-1833 (Electronic))

3. Darst BF, Sheng X, Eeles RA, Kote-Jarai Z, Conti DV, Haiman CA. Combined Effect of a Polygenic Risk Score and Rare Genetic Variants on Prostate Cancer Risk. (1873-7560 (Electronic))

4. Conti DV, Darst BA-O, Moss LC, et al. Trans-ancestry genome-wide association meta-analysis of prostate cancer identifies new susceptibility loci and informs genetic risk prediction. (1546-1718 (Electronic))

5. Huynh-Le MA-O, Karunamuni R, Fan CA-O, et al. Prostate cancer risk stratification improvement across multiple ancestries with new polygenic hazard score. LID - 10.1038/s41391-022-00497-7 [doi]. (1476-5608 (Electronic))

6. Huynh-Le MA-O, Myklebust TÅ A-O, Feng CA-O, et al. Age dependence of modern clinical risk groups for localized prostate cancer-A population-based study. (1097-0142 (Electronic))

7. Chinea FM, Patel VN, Kwon D, et al. Ethnic heterogeneity and prostate cancer mortality in Hispanic/Latino men: a population-based study. Oncotarget. Sep 19 2017;8(41):69709–69721. doi:10.18632/oncotarget.19068

8. Wilson KM, Mucci LA. Diet and Lifestyle in Prostate Cancer. (0065-2598 (Print))

9. Rivera-Izquierdo MA-O, Pérez de Rojas J, Martínez-Ruiz V, Arrabal-Polo M, Pérez-Gómez BA-O, Jiménez-Moleón JA-O. Obesity and biochemical recurrence in clinically localised prostate cancer: a systematic review and meta-analysis of 86,490 patients. LID - 10.1038/s41391-021-00481-7 [doi]. (1476-5608 (Electronic))

10. Zhang X, Zhou G, Sun B, et al. Impact of obesity upon prostate cancer-associated mortality: A meta-analysis of 17 cohort studies. (1792-1074 (Print))

11. Islami F, Moreira DM, Boffetta P, Freedland SJ. A systematic review and meta-analysis of tobacco use and prostate cancer mortality and incidence in prospective cohort studies. (1873-7560 (Electronic))

12. Langlais CS, Graff RE, Van Blarigan EL, et al. Post-Diagnostic Dietary and Lifestyle Factors and Prostate Cancer Recurrence, Progression, and Mortality. Current oncology reports. 2021;23(3):37–37. doi:10.1007/s11912-021-01017-x

13. Key TA-OX, Appleby PN, Travis RC, et al. Carotenoids, retinol, tocopherols, and prostate cancer risk: pooled analysis of 15 studies. (1938-3207 (Electronic))

14. Plym A, Zhang Y, Stopsack KH, et al. A Healthy Lifestyle in Men at Increased Genetic Risk for Prostate Cancer. Eur Urol. May 27 2022;doi:10.1016/j.eururo.2022.05.008

15. Khera AV, Emdin CA, Drake I, et al. Genetic Risk, Adherence to a Healthy Lifestyle, and Coronary Disease. (1533-4406 (Electronic))

16. Arthur RS, Wang T, Xue X, Kamensky V, Rohan TE. Genetic Factors, Adherence to Healthy Lifestyle Behavior, and Risk of Invasive Breast Cancer Among Women in the UK Biobank. JNCI: Journal of the National Cancer Institute. 2020;112(9):893–901. doi:10.1093/jnci/djz241

17. Choi J, Jia G, Wen W, Shu XO, Zheng W. Healthy lifestyles, genetic modifiers, and colorectal cancer risk: a prospective cohort study in the UK Biobank. (1938-3207 (Electronic))

18. Gaziano JM, Concato J, Brophy M, et al. Million Veteran Program: A mega-biobank to study genetic influences on health and disease. (1878-5921 (Electronic))

19. Pagadala M, Lynch JA, Karunamuni R, et al. Evaluating a polygenic hazard score to predict risk of developing metastatic or fatal prostate cancer in the multi-ancestry Million Veteran Program cohort. Journal of Clinical Oncology. 2022;40(6_suppl):155–155. doi:10.1200/JCO.2022.40.6_suppl.155

20. Hunter-Zinck H, Shi Y, Li M, et al. Genotyping Array Design and Data Quality Control in the Million Veteran Program. (1537-6605 (Electronic))

21. Hugosson J, Roobol MJ, Månsson M, et al. A 16-yr Follow-up of the European Randomized study of Screening for Prostate Cancer. Eur Urol. 07 2019;76(1):43–51. doi:10.1016/j.eururo.2019.02.009

22. Flegal KM, Graubard BI. Estimates of excess deaths associated with body mass index and other anthropometric variables. (1938-3207 (Electronic))

23. North American Association for the Study of Obesity National Heart Lung and Blood Institute National Institutes of Health (U.S.) NHLBI Obesity Education Initiative. The Practical Guide : Identification Evaluation and Treatment of Overweight and Obesity in Adults.. Bethesda Md: National Institutes of Health National Heart Lung and Blood Institute NHLBI Obesity Education Initiative North American Association for the Study of Obesity; 2000.

24. Rebbeck TR. Prostate Cancer Disparities by Race and Ethnicity: From Nucleotide to Neighborhood. LID - 10.1101/cshperspect.a030387 [doi] LID - a030387. (2157-1422 (Electronic))

25. Graham-Steed T, Uchio E Fau - Wells CK, Wells Ck Fau - Aslan M, Aslan M Fau - Ko J, Ko J Fau - Concato J, Concato J. ‘Race’ and prostate cancer mortality in equal-access healthcare systems. (1555-7162 (Electronic))

26. Coughlin SS. A review of social determinants of prostate cancer risk, stage, and survival. (2287-8882 (Print))

27. Zavala VA, Bracci PM, Carethers JM, et al. Cancer health disparities in racial/ethnic minorities in the United States. (1532-1827 (Electronic))

28. Stern MC, Fejerman L, Das R, et al. Variability in Cancer Risk and Outcomes Within US Latinos by National Origin and Genetic Ancestry. (2196-2995 (Print))

29. Leal-García M, Canto P, Cárdenas-Cárdenas E, Feria-Bernal G, García-García E, Méndez JP. Overweight and obesity in men with prostate cancer do not constitute risk factors for biochemical recurrence. (1473-0790 (Electronic))

30. Martini AA-O, Shah QN, Waingankar N, et al. The obesity paradox in metastatic castration-resistant prostate cancer. (1476-5608 (Electronic))

31. Freedland Sj Fau - Aronson WJ, Aronson WJ. Examining the relationship between obesity and prostate cancer. (1523-6161 (Print))

