## SupplementaryTable 1 for "Healthy lifestyle and prostate cancer risk in the Million Veteran Program"

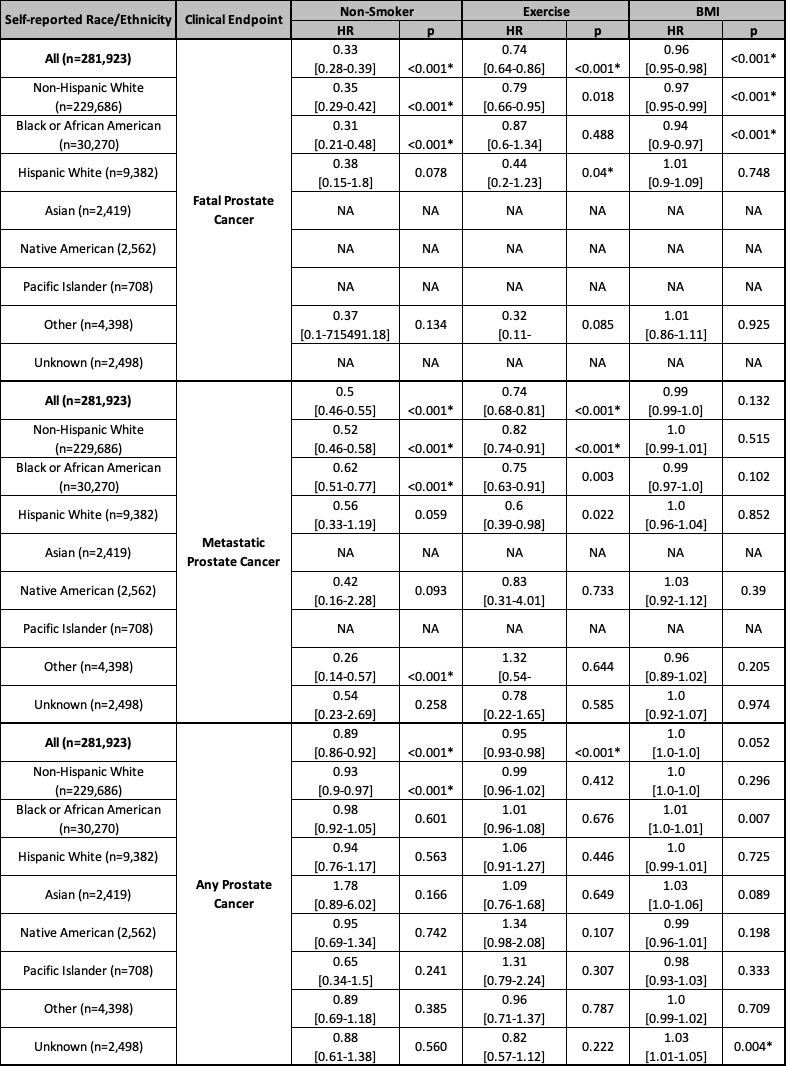

**Supplementary Table 1: Univariable association of lifestyle factors with any, metastatic and fatal prostate cancer, stratified by racial/ethnic group.** Cox proportional hazards model results from association with age at prostate cancer, metastatic prostate cancer (nodal or distant) and death from prostate cancer. *P*-values reported are from univariable models using each lifestyle factor, respectively, as the sole predictor variable. The hazard ratio (HR) for smoking status is for non-smokers, (reference is smokers). The HR for exercise is for those participating in vigorous activity 2 or more days per week (reference is exercise fewer than 2 days per week). The HR for BMI (continuous variable) is per kg/m^2^. Significant predictors are indicated by *. NA, indicates subgroup analyses limited by sample size. n, indicates number at risk.
