## Supplementary Table 2 for "Healthy lifestyle and prostate cancer risk in the Million Veteran Program"

|  | **Race/Ethnicity** | | | | | | | **FH** | **Healthy Lifestyle** | | |
| --- | --- | --- | --- | --- | --- | --- | --- | --- | --- | --- | --- |
| **Clinical Endpoint** | **Black** | **Hispanic** | **Asian** | **Native American** | **Pacific Islander** | **Other** | **Unknown** |  | **Non-smoker** | **BMI** | **Exercise** |
| Fatal Prostate Cancer | 2.41  [1.93-2.94]*** | 0.96  [0.52-1.48] | 0.23  [0.0-0.7] | 1.15  [0.27-2.48] | 1.72  [0.0-6.06] | 1.51  [0.56-2.69] | 0.0  [0.0-0.0] | 2.02  [1.61-2.46]*** | 0.36  [0.3-0.43]*** | 0.96  [0.94-0.98]** | 0.84  [0.68-1.07] |
| Metastatic Prostate Cancer | 2.76  [2.48-3.08]*** | 1.21  [0.92-1.53] | 0.88  [0.38-1.36] | 1.01  [0.5-1.64] | 0.97  [0.0-2.59] | 1.68  [1.09-2.31]* | 1.71  [0.66-2.92] | 1.72  [1.49-1.95]*** | 0.52  [0.47-0.58]*** | 0.99  [0.98-1.0] | 0.89  [0.77-1.03] |
| Prostate Cancer | 2.3  [2.21-2.38]*** | 1.08  [1.0-1.17] | 0.92  [0.79-1.07] | 1.06  [0.91-1.22] | 1.17  [0.83-1.56] | 1.12  [1.0-1.24] | 0.86  [0.65-1.13] | 1.93  [1.86-1.99]*** | 0.92  [0.89-0.95]** | 1.01  [1.0-1.01]** | 0.99  [0.95-1.04] |

**Supplementary Table 2: Multivariable models combining self-reported Race/Ethnicity, Family History (FH) and lifestyle factors for only participants with BMI between 18.5 and 35.** Cox proportional hazards results for association with age at death from prostate cancer, at diagnosis of metastatic prostate cancer and age at diagnosis with prostate cancer for participants with BMI 18.5-35. *P*-values reported are from multivariable models using self-reported race/ethnicity, family history and three lifestyle factors (non-smoker, BMI, and exercise). Hazard ratios for race/ethnicity were estimated using Non-Hispanic White as reference. Hazard ratios for family history were for one or more first-degree relatives diagnosed with prostate cancer. Numbers in brackets are 95% confidence intervals. Significant predictors in the multivariable model are indicated by *(*p*<0.01), **(p<10^-6^) and *** (p<10^-16^).
