## Supplementary Table 3 for "Healthy lifestyle and prostate cancer risk in the Million Veteran Program"

| **Intercept** | **Race/Ethnicity** | | | | | | | **FH** | **Age** | **Lifestyle** | | |
| --- | --- | --- | --- | --- | --- | --- | --- | --- | --- | --- | --- | --- |
|  | **Black** | **Hispanic** | **Asian** | **Native American** | **Pacific Islander** | **Other** | **Unknown** |  |  | **Non-Smoker** | **BMI** | **Exercise** |
| -2.48 | 1.4 | -0.33 | -1.1 | 1.17 | -0.18 | 0.69 | -2.04 | 0.11 | 0.12 | -1.05 | 0.10 | -0.24 |
| -[2.74-2.22]** | [1.31-1.49]** | -[0.49-0.64]** | -[1.39-0.82]** | [0.90-1.44]** | [-0.84-0.47] | [0.49-0.89]** | -[2.59-1.49]** | [0.01-0.21] | [0.11-0.12] | -[1.12-0.97]** | [0.09-0.10]** | -[0.34-0.14]** |

**Supplementary Table 3: Multivariable linear regression assessing association between self-reported Race/Ethnicity, Family History (FH), and lifestyle factors with screening intensity (total number of PSA screening tests) among participants without PCa.** Results for association between each factor and number of PSA tests recorded in the VA medical record for a given participant. *P*-values reported are from linear model using self-reported race/ethnicity, family history, and three lifestyle factors (non-smoker, BMI, and exercise). Coefficients for race/ethnicity were estimated using Non-Hispanic White as reference. Family history was defined as one or more first-degree relatives diagnosed with prostate cancer. Coefficient for Non-Smoker used Smoker as reference—i.e., non-smokers underwent, on average, one less PSA test. Coefficient for BMI is per unit of BMI. Coefficient for exercise was for those who reported vigorous exercise ≥2 times per week, with exercise <2 times per week as the reference. Numbers in brackets are 95% confidence intervals. Significant predictors in the multivariable model are indicated by *(*p*<0.01), **(p<0.001).
