## Supplementary Table 4 for "Healthy lifestyle and prostate cancer risk in the Million Veteran Program"

| **Variable** | **Description** | **Any Prostate Cancer** | | **Metastatic Prostate Cancer** | | **Fatal Prostate Cancer** | |
| --- | --- | --- | --- | --- | --- | --- | --- |
|  |  | **HR** | **p-value** | **HR** | **p-value** | **HR** | **p-value** |
| alcfreq | How often a drink that contains alcohol? | 1.01 [1.01-1.02] | 3.14E-07 | 0.97 [0.95-0.99] | 4.70E-04 | 0.96 [0.93-1.0] | 6.24E-03 |
| alcqty | How many drinks containing alcohol on a typical day when you are drinking? | 1.08 [1.06-1.11] | 2.64E-25 | 1.35 [1.26-1.43] | 1.61E-33 | 1.36 [1.18-1.53] | 1.20E-09 |
| bakebutter | Subject uses real butter for baking at home | 1.02 [1.0-1.04] | 1.18E-02 | 1.04 [0.97-1.12] | 2.43E-01 | 0.94 [0.82-1.06] | 3.45E-01 |
| bakelard | Subject uses lard for baking at home | 0.95 [0.87-1.04] | 1.31E-01 | 1.05 [0.72-1.38] | 7.62E-01 | 0.91 [0.44-1.43] | 7.46E-01 |
| bakemarg | Subject uses margarine for baking at home | 1.03 [1.0-1.05] | 2.88E-03 | 0.93 [0.85-1.01] | 9.02E-02 | 0.99 [0.84-1.15] | 8.78E-01 |
| bakevegoil | Subject uses vegetable oil for baking at home | 1.0 [0.98-1.02] | 8.22E-01 | 1.05 [0.98-1.13] | 1.65E-01 | 1.11 [0.97-1.25] | 9.65E-02 |
| bakevegshort | Subject uses vegetable shortening for baking at home | 0.98 [0.95-1.01] | 6.11E-02 | 0.88 [0.78-0.99] | 4.38E-02 | 0.89 [0.71-1.07] | 2.67E-01 |
| circcad | Have you been diagnosed with Coronary Artery / Coronary Heart Disease (including heart attack, angina) | 0.78 [0.76-0.81] | 1.50E-131 | 0.65 [0.59-0.72] | 2.72E-32 | 0.75 [0.63-0.88] | 1.25E-06 |
| circcadmed | Do you take medication for Coronary Artery / Coronary Heart Disease | 0.79 [0.76-0.81] | 1.40E-111 | 0.65 [0.59-0.73] | 9.49E-28 | 0.74 [0.62-0.87] | 3.61E-06 |
| circchf | Have you been diagnosed with Congestive Heart Failure | 0.78 [0.75-0.81] | 7.71E-55 | 0.64 [0.54-0.74] | 6.03E-14 | 0.64 [0.49-0.82] | 1.58E-05 |
| circchfmed | Do you take medication for High blood pressure / hypertension | 0.79 [0.75-0.83] | 5.57E-43 | 0.68 [0.57-0.79] | 6.84E-10 | 0.64 [0.47-0.85] | 5.55E-05 |
| circchol | Have you been diagnosed with High Cholesterol | 1.03 [1.0-1.05] | 1.22E-03 | 0.88 [0.82-0.94] | 5.63E-07 | 0.96 [0.84-1.09] | 4.05E-01 |
| circcholmed | Do you take medication for High Cholesterol | 1.01 [0.99-1.03] | 3.27E-01 | 0.86 [0.8-0.92] | 8.67E-09 | 0.91 [0.8-1.03] | 4.90E-02 |
| circhrtatk | Have you been diagnosed with Heart Attack | 0.81 [0.79-0.84] | 9.48E-74 | 0.72 [0.64-0.8] | 1.88E-16 | 0.92 [0.77-1.07] | 1.82E-01 |
| circhrtatkmed | Do you take medication for Heart Attack | 0.81 [0.78-0.83] | 1.10E-69 | 0.76 [0.67-0.84] | 4.46E-11 | 0.82 [0.67-0.98] | 4.13E-03 |
| circhtn | Have you been diagnosed with High blood pressure / hypertension | 1.02 [1.0-1.04] | 3.29E-03 | 0.97 [0.9-1.04] | 2.94E-01 | 0.99 [0.88-1.14] | 8.86E-01 |
| circhtnmed | Do you take medication for High blood pressure / hypertension | 1.01 [0.98-1.03] | 3.19E-01 | 0.94 [0.87-1.01] | 1.51E-02 | 0.95 [0.84-1.09] | 2.63E-01 |
| circpvd | Have you been diagnosed with Peripheral Vascular Disease | 0.84 [0.8-0.88] | 1.81E-23 | 0.76 [0.63-0.88] | 1.48E-05 | 0.87 [0.66-1.14] | 1.74E-01 |
| circpvdmed | Do you take medication for Peripheral Vascular Disease | 0.83 [0.78-0.87] | 4.36E-20 | 0.79 [0.63-0.94] | 1.13E-03 | 0.99 [0.69-1.32] | 9.48E-01 |
| circstrk | Have you been diagnosed with Stroke | 0.94 [0.9-0.98] | 4.19E-05 | 0.84 [0.71-0.96] | 7.49E-04 | 0.9 [0.69-1.13] | 2.23E-01 |
| circstrkmed | Do you take medication for Stroke | 0.92 [0.88-0.97] | 4.03E-06 | 0.9 [0.76-1.05] | 8.27E-02 | 1.03 [0.76-1.31] | 7.82E-01 |
| circtia | Have you been diagnosed with Transient Ischemic Attack (TIA) | 0.92 [0.88-0.97] | 5.64E-06 | 0.58 [0.47-0.69] | 2.26E-14 | 0.69 [0.49-0.9] | 6.84E-04 |
| circtiamed | Do you take medication for Transient Ischemic Attack (TIA) | 0.93 [0.87-0.98] | 2.45E-04 | 0.71 [0.56-0.86] | 2.02E-05 | 0.88 [0.62-1.19] | 3.14E-01 |
| dodm | Have you been diagnosed with Diabetes/Sugar | 0.87 [0.85-0.89] | 1.44E-57 | 0.91 [0.84-0.99] | 2.06E-03 | 1.05 [0.91-1.2] | 3.72E-01 |
| dodmmed | Do you take medication for Diabetes/ Sugar | 0.85 [0.83-0.87] | 3.04E-70 | 0.9 [0.83-0.98] | 1.43E-03 | 1.03 [0.89-1.19] | 5.53E-01 |
| ffqbacon | Subject's average consumption of bacon (2 slices), in the last year | 1.02 [1.01-1.03] | 9.61E-10 | 1.04 [1.02-1.07] | 2.00E-05 | 1.04 [0.99-1.09] | 2.62E-02 |
| ffqbeef | Subject's average consumption of Beef, pork, or lamb as a sandwich or mixed dish, in the last year | 1.0 [0.99-1.01] | 9.06E-01 | 1.02 [0.99-1.05] | 1.25E-01 | 1.06 [1.0-1.12] | 3.81E-03 |
| ffqbeef_2 | Subject's average consumption of beef, pork, or lamb as a main dish (4-6 oz) in the last year | 1.0 [0.99-1.01] | 5.06E-01 | 1.0 [0.97-1.03] | 9.67E-01 | 1.03 [0.97-1.09] | 1.84E-01 |
| ffqbeer | Subject's average consumption of beer (1 glass, bottle, can) in the last year | 1.02 [1.01-1.02] | 7.28E-17 | 1.0 [0.97-1.02] | 7.01E-01 | 0.94 [0.9-0.98] | 2.72E-04 |
| ffqcake | Subject's average consumption of cake (slice) in the last year | 0.99 [0.98-1.0] | 8.73E-02 | 0.93 [0.89-0.96] | 3.29E-07 | 0.97 [0.91-1.04] | 2.83E-01 |
| ffqcandy | Subject's average consumption of candy without chocolate (1 oz.) in the last year | 1.0 [0.99-1.0] | 9.65E-02 | 0.97 [0.94-0.99] | 4.76E-04 | 0.99 [0.94-1.03] | 4.30E-01 |
| ffqcarbbev | Subject's average consumption of carbonated beverages with sugar in the last year | 1.03 [1.02-1.03] | 4.19E-26 | 1.03 [1.01-1.06] | 3.60E-05 | 1.05 [1.0-1.09] | 3.31E-03 |
| ffqchips | Subject's average consumption of potato chips or corn chips (small bag or 1 oz.) in the last year | 1.01 [1.0-1.01] | 3.58E-02 | 1.0 [0.97-1.02] | 7.18E-01 | 0.98 [0.94-1.03] | 3.80E-01 |
| ffqchknoskn | Subject's average consumption of chicken or turkey, without skin (4 - 6 oz), in the last year | 1.01 [1.0-1.02] | 6.22E-02 | 0.99 [0.96-1.02] | 2.32E-01 | 1.02 [0.96-1.08] | 3.80E-01 |
| ffqchoc | Subject's average consumption of chocolate (1 oz.) in the last year | 0.98 [0.97-0.99] | 2.09E-19 | 0.93 [0.91-0.96] | 3.90E-15 | 0.96 [0.92-0.99] | 2.25E-03 |
| ffqcknskn | Subject's average consumption of chicken or turkey, with skin (4 - 6 oz), in the last year | 1.04 [1.03-1.05] | 1.10E-44 | 1.08 [1.06-1.11] | 1.17E-16 | 1.05 [1.0-1.1] | 3.22E-03 |
| ffqcldcer | Subject's average consumption of cold breakfast cereal (1 cup) in the last year | 0.98 [0.98-0.99] | 4.25E-13 | 0.93 [0.91-0.95] | 9.40E-22 | 0.98 [0.95-1.02] | 2.09E-01 |
| ffqcof | Subject's average consumption of caffeinated coffee (1 cup) in the last year | 0.98 [0.98-0.98] | 1.26E-47 | 0.99 [0.98-1.0] | 2.91E-02 | 0.96 [0.94-0.98] | 5.15E-06 |
| ffqcookie | Subject's average consumption of cookies (1) in the last year | 0.98 [0.97-0.98] | 2.52E-22 | 0.9 [0.88-0.92] | 6.70E-32 | 0.92 [0.88-0.96] | 3.52E-08 |
| ffqdbttr | Subject's average consumption of butter (pat), added to food or bread, in the last year | 1.0 [0.99-1.0] | 6.46E-02 | 1.0 [0.99-1.02] | 4.89E-01 | 1.0 [0.97-1.03] | 7.57E-01 |
| ffqdcottage | Subject's average consumption of cottage or ricotta cheese (1/2 cup) in the last year | 0.97 [0.96-0.98] | 7.76E-22 | 0.99 [0.96-1.02] | 2.76E-01 | 1.02 [0.96-1.08] | 2.42E-01 |
| ffqdicecrm | Subject's average consumption of ice cream (1 cup) in the last year | 0.97 [0.96-0.98] | 8.39E-25 | 0.93 [0.91-0.96] | 3.88E-13 | 1.02 [0.98-1.07] | 2.19E-01 |
| ffqdmarg | Subject's average consumption of margarine (pat), added to food or bread, in the last year | 0.99 [0.99-1.0] | 1.75E-03 | 0.99 [0.97-1.01] | 8.16E-02 | 1.01 [0.98-1.04] | 2.71E-01 |
| ffqdothchs | Subject's average consumption of other cheese (1 slice or 1 oz. serving) in the last year | 0.98 [0.97-0.99] | 5.19E-14 | 0.96 [0.94-0.99] | 2.74E-05 | 0.98 [0.94-1.03] | 2.50E-01 |
| ffqdressing | Subject's average consumption of other oil and vinegar dressing, e.g., Italian (1 Tbs) in the last year | 1.0 [0.99-1.01] | 9.12E-01 | 0.96 [0.93-0.98] | 6.94E-07 | 0.96 [0.92-1.0] | 1.79E-02 |
| ffqdrkbrd | Subject's average consumption of dark bread, including wheat pita bread (slice), in the last year | 1.0 [0.99-1.0] | 6.36E-02 | 0.99 [0.97-1.01] | 9.21E-02 | 0.98 [0.95-1.02] | 1.95E-01 |
| ffqdskim | Subject's average consumption of skim or low fat milk (8 oz. glass) in the last year | 0.99 [0.98-0.99] | 2.24E-11 | 0.95 [0.93-0.97] | 2.06E-18 | 0.99 [0.96-1.01] | 1.47E-01 |
| ffqdtcarbbev | Subject's average consumption of low calorie carbonated beverages in the last year | 0.97 [0.97-0.98] | 2.74E-36 | 0.94 [0.92-0.97] | 7.39E-14 | 0.98 [0.95-1.02] | 2.33E-01 |
| ffqdwhl | Subject's average consumption of whole milk (8 oz. glass) in the last year | 1.0 [1.0-1.01] | 2.29E-01 | 1.04 [1.01-1.06] | 5.04E-06 | 1.04 [1.0-1.08] | 4.34E-03 |
| ffqdyog | Subject's average consumption of yogurt (1 cup) in the last year | 1.01 [1.0-1.01] | 1.93E-02 | 0.98 [0.96-1.0] | 1.67E-02 | 1.01 [0.97-1.05] | 5.11E-01 |
| ffqegg | Subject's average consumption of eggs (1) in the last year | 0.99 [0.99-1.0] | 4.87E-03 | 1.02 [1.0-1.05] | 1.56E-02 | 1.07 [1.03-1.12] | 1.42E-05 |
| ffqfappl | Subject's average consumption of fresh apples or pears, in the last year | 1.0 [0.99-1.01] | 8.77E-01 | 0.99 [0.96-1.01] | 1.07E-01 | 1.0 [0.96-1.05] | 8.95E-01 |
| ffqfat | Amount of visible fat on beef/pork/lamb that subject removes before eating | 1.02 [1.01-1.03] | 6.59E-06 | 1.08 [1.04-1.11] | 1.15E-10 | 0.99 [0.94-1.06] | 7.39E-01 |
| ffqfbanan | Subject's average consumption of bananas in the last year | 0.99 [0.99-1.0] | 8.96E-04 | 0.96 [0.94-0.98] | 7.33E-08 | 0.98 [0.95-1.02] | 2.47E-01 |
| ffqfish | Subject's average consumption of fish (3-5 oz.) in the last year | 1.03 [1.02-1.04] | 1.32E-16 | 1.01 [0.98-1.05] | 3.42E-01 | 0.95 [0.89-1.0] | 1.27E-02 |
| ffqfojgfj | Subject's average consumption of orange juice or grapefruit juice (small glass), in the last year | 1.0 [0.99-1.0] | 7.61E-02 | 0.97 [0.95-0.99] | 1.86E-05 | 0.96 [0.93-0.99] | 6.23E-04 |
| ffqforng | Subject's average consumption of oranges, in the last year | 1.0 [1.0-1.01] | 1.53E-01 | 1.01 [0.98-1.03] | 3.55E-01 | 1.01 [0.97-1.06] | 6.52E-01 |
| ffqfoth | Subject's average consumption of other fruits (fresh, frozen, or canned) in the last year | 0.99 [0.99-1.0] | 3.51E-03 | 0.98 [0.95-1.0] | 4.62E-03 | 1.02 [0.99-1.06] | 1.20E-01 |
| ffqfpeach | Subject's average consumption of peaches, apricots, or plums (1 fresh, or 1/2 cup canned), in the last year | 0.99 [0.98-1.0] | 9.42E-06 | 0.97 [0.95-1.0] | 1.01E-02 | 1.01 [0.97-1.06] | 3.93E-01 |
| ffqfrudrnk | Subject's average consumption of Hawaiian Punch, lemonade, or other fruit drinks in the last year | 1.03 [1.02-1.04] | 3.64E-33 | 1.07 [1.05-1.09] | 7.23E-16 | 1.06 [1.02-1.11] | 3.56E-05 |
| ffqfryaway | Subject's consumption of fried foods away from home | 1.02 [1.0-1.04] | 2.41E-03 | 1.01 [0.95-1.08] | 5.57E-01 | 1.04 [0.92-1.17] | 3.41E-01 |
| ffqfryhome | Subject's consumption of fried foods at home | 0.98 [0.97-1.0] | 1.32E-03 | 1.06 [1.01-1.11] | 1.43E-03 | 1.08 [0.97-1.18] | 2.29E-02 |
| ffqhambrg | Subject's average consumption of hamburger (1 patty), in the last year | 0.97 [0.96-0.98] | 1.59E-13 | 0.97 [0.94-1.01] | 3.41E-02 | 1.06 [0.99-1.13] | 7.74E-03 |
| ffqhtdog | Subject's average consumption of hot dogs (1), in the last year | 1.0 [0.99-1.01] | 9.07E-01 | 1.03 [0.99-1.07] | 2.76E-02 | 1.08 [1.01-1.15] | 8.47E-04 |
| ffqliquor | Subject's average consumption of liquor (1 drink or shot) in the last year | 1.01 [1.01-1.02] | 7.02E-10 | 1.0 [0.97-1.02] | 6.74E-01 | 0.99 [0.95-1.03] | 6.46E-01 |
| ffqliver | Subject's average consumption of liver (3-4 oz), in the last year | 1.05 [1.03-1.07] | 1.07E-14 | 1.15 [1.09-1.22] | 4.63E-11 | 1.08 [0.96-1.2] | 7.36E-02 |
| ffqnuts | Subject's average consumption of nuts (small packet or 1 oz.) in the last year | 1.0 [0.99-1.0] | 2.38E-01 | 0.96 [0.93-0.98] | 1.17E-07 | 0.94 [0.9-0.98] | 5.05E-05 |
| ffqpb | Subject's average consumption of peanut butter (1 Tbs) in the last year | 0.97 [0.97-0.98] | 2.21E-28 | 0.97 [0.95-1.0] | 7.41E-04 | 0.99 [0.95-1.03] | 4.27E-01 |
| ffqpiehm | Subject's average consumption of homemade pie (slice) in the last year | 1.0 [0.99-1.01] | 4.89E-01 | 0.97 [0.93-1.01] | 3.91E-02 | 1.01 [0.94-1.09] | 6.88E-01 |
| ffqpierm | Subject's average consumption of ready made pie (slice) in the last year | 0.99 [0.98-1.01] | 1.86E-01 | 0.96 [0.92-1.0] | 7.56E-03 | 0.99 [0.92-1.06] | 7.77E-01 |
| ffqpotbkd | Subject's average consumption of baked, boiled (1) or mashed (1 cup) potatoes in the last year | 0.98 [0.97-0.99] | 2.93E-10 | 0.97 [0.93-1.0] | 1.74E-03 | 1.0 [0.95-1.06] | 8.41E-01 |
| ffqpotfried | Subject's average consumption of French fried potatoes (4 oz.) in the last year | 1.02 [1.01-1.03] | 1.08E-06 | 1.03 [1.0-1.07] | 6.02E-03 | 1.04 [0.98-1.11] | 4.95E-02 |
| ffqprocmeat | Subject's average consumption of processed meats e.g., sausage, salami, bologna, etc (piece or slice), in the last year | 0.99 [0.98-1.0] | 8.27E-04 | 0.97 [0.95-1.0] | 1.06E-02 | 1.01 [0.96-1.06] | 5.50E-01 |
| ffqrice | Subject's average consumption of rice or pasta, e.g., spaghetti, noodles, etc. (1 cup) in the last year | 1.01 [1.0-1.02] | 1.13E-04 | 1.03 [1.0-1.07] | 4.49E-03 | 1.07 [1.01-1.13] | 8.02E-04 |
| ffqrxdiet | Subject follows a physician-prescribed special diet | 0.97 [0.95-1.0] | 2.31E-03 | 0.93 [0.85-1.01] | 1.69E-02 | 1.03 [0.89-1.2] | 5.51E-01 |
| ffqrxdietchol | Subject follows a low cholesterol diet | 1.0 [0.97-1.04] | 6.90E-01 | 0.97 [0.86-1.07] | 4.32E-01 | 0.99 [0.8-1.2] | 8.67E-01 |
| ffqrxdietdiab | Subject follows a diabetic diet | 0.92 [0.89-0.95] | 1.10E-13 | 0.89 [0.79-1.0] | 6.80E-03 | 0.98 [0.79-1.2] | 7.46E-01 |
| ffqrxdietfat | Subject follows a low-fat diet | 0.94 [0.9-0.98] | 2.42E-05 | 0.87 [0.75-0.99] | 6.13E-03 | 0.96 [0.76-1.21] | 6.70E-01 |
| ffqrxdietoth | Subject follows another physician-prescribed diet | 1.03 [0.96-1.09] | 2.17E-01 | 0.89 [0.69-1.11] | 1.66E-01 | 1.19 [0.8-1.65] | 1.68E-01 |
| ffqrxdietpot | Subject follows a high potassium diet | 1.02 [0.93-1.11] | 5.60E-01 | 1.03 [0.75-1.33] | 7.61E-01 | 1.2 [0.74-1.8] | 2.63E-01 |
| ffqrxdietsod | Subject follows a low sodium diet | 0.97 [0.93-1.0] | 4.77E-03 | 0.88 [0.78-0.99] | 4.47E-03 | 0.92 [0.74-1.12] | 2.57E-01 |
| ffqrxdiettrig | Subject follows a low triglyceride diet | 0.92 [0.88-0.98] | 2.06E-04 | 0.79 [0.61-0.97] | 2.63E-03 | 0.79 [0.51-1.11] | 1.04E-01 |
| ffqrxdietulc | Subject follows an ulcer diet | 1.03 [0.9-1.19] | 4.93E-01 | 0.82 [0.45-1.25] | 2.74E-01 | 1.41 [0.59-2.36] | 1.46E-01 |
| ffqrxdietwtred | Subject follows a weight reduction (low calorie) diet | 1.06 [1.02-1.11] | 8.22E-05 | 1.0 [0.85-1.15] | 9.57E-01 | 1.02 [0.73-1.31] | 8.66E-01 |
| ffqsugar | Teaspoons of sugar that subject adds to beverages or food daily | 1.0 [1.0-1.0] | 4.84E-08 | 1.0 [1.0-1.0] | 2.39E-03 | 1.0 [0.99-1.01] | 1.77E-04 |
| ffqvbean | Subject's average consumption of baked or dried beans or lentils (1/2 cup) in the last year | 0.99 [0.99-1.0] | 7.29E-02 | 1.01 [0.98-1.04] | 2.36E-01 | 1.0 [0.95-1.06] | 9.32E-01 |
| ffqvbroc | Subject's average consumption of broccoli (1/2 cup) in the last year | 1.03 [1.02-1.03] | 3.62E-16 | 1.05 [1.02-1.08] | 8.38E-06 | 1.03 [0.97-1.09] | 1.34E-01 |
| ffqvcarcook | Subject's average consumption of cooked carrots (1/2 carrot or 2-4 sticks) in the last year | 0.99 [0.98-1.0] | 3.56E-04 | 1.01 [0.98-1.04] | 2.85E-01 | 1.02 [0.96-1.08] | 3.72E-01 |
| ffqvcarraw | Subject's average consumption of raw carrots (1/2 carrot or 2-4 sticks) in the last year | 1.0 [1.0-1.01] | 1.11E-01 | 1.0 [0.97-1.03] | 9.16E-01 | 0.96 [0.92-1.02] | 3.66E-02 |
| ffqvcbg | Subject's average consumption of cabbage, cauliflower or Brussels sprouts (1/2 cup) in the last year | 1.01 [1.0-1.02] | 4.83E-02 | 1.05 [1.02-1.08] | 3.17E-06 | 1.06 [1.0-1.12] | 3.16E-03 |
| ffqvcorn | Subject's average consumption of corn (1 ear or 1/2 cup frozen or canned) in the last year | 1.0 [0.99-1.01] | 4.18E-01 | 1.0 [0.97-1.04] | 7.40E-01 | 1.0 [0.95-1.06] | 9.08E-01 |
| ffqvpea | Subject's average consumption of peas or lima beans (1/2 cup fresh, frozen canned) in the last year | 1.0 [0.99-1.01] | 5.62E-01 | 1.0 [0.97-1.04] | 6.92E-01 | 1.0 [0.94-1.06] | 8.33E-01 |
| ffqvspin | Subject's average consumption of cooked spinach or collard greens (1/2cup) in the last year | 1.04 [1.04-1.05] | 2.80E-44 | 1.08 [1.05-1.12] | 2.27E-14 | 1.05 [1.0-1.11] | 1.07E-02 |
| ffqvsqsh | Subject's average consumption of yellow (winter) squash (1/2 cup) in the last year | 1.0 [0.99-1.01] | 4.94E-01 | 0.98 [0.95-1.02] | 2.76E-01 | 1.02 [0.95-1.1] | 3.74E-01 |
| ffqvstngbns | Subject's average consumption of string beans (1/2 cup) in the last year | 1.01 [1.0-1.02] | 5.04E-02 | 0.99 [0.96-1.02] | 5.17E-01 | 0.99 [0.93-1.04] | 6.00E-01 |
| ffqvtom | Subject's average consumption of tomatoes (1) or tomato juice (small glass) in the last year | 0.98 [0.97-0.98] | 2.03E-20 | 0.99 [0.96-1.01] | 1.38E-01 | 0.97 [0.93-1.0] | 1.76E-02 |
| ffqvyam | Subject's average consumption of yams or sweet potatoes (1/2 cup) in the last year | 1.02 [1.01-1.03] | 1.93E-08 | 1.0 [0.97-1.04] | 7.55E-01 | 1.01 [0.95-1.07] | 7.12E-01 |
| ffqwhitbrd | Subject's average consumption of white bread, including pita bread (slice), in the last year | 0.99 [0.99-1.0] | 1.93E-05 | 0.99 [0.97-1.01] | 5.12E-02 | 0.99 [0.96-1.03] | 5.36E-01 |
| frybutter | Subject uses real butter for frying or sautéing at home | 0.98 [0.96-1.0] | 1.35E-02 | 1.01 [0.94-1.09] | 7.29E-01 | 0.91 [0.78-1.05] | 7.58E-02 |
| frylard | Subject uses lard for frying or sautéing at home | 0.92 [0.84-1.02] | 1.99E-02 | 1.09 [0.78-1.47] | 4.05E-01 | 1.06 [0.47-1.71] | 7.68E-01 |
| frymarg | Subject uses real butter for frying or sautéing at home | 1.01 [0.98-1.04] | 2.63E-01 | 0.94 [0.85-1.03] | 7.56E-02 | 1.06 [0.9-1.24] | 2.91E-01 |
| fryvegoil | Subject uses margarine for frying or sautéing at home | 1.03 [1.01-1.06] | 2.57E-05 | 1.05 [0.98-1.14] | 5.10E-02 | 1.06 [0.94-1.22] | 1.96E-01 |
| fryvegshort | Subject uses vegetable oil for frying or sautéing at home | 1.0 [0.96-1.05] | 7.80E-01 | 0.9 [0.76-1.05] | 6.19E-02 | 0.95 [0.7-1.21] | 6.03E-01 |
| vita | Subject regularly takes Vitamin A | 0.97 [0.93-1.01] | 6.06E-02 | 1.05 [0.92-1.19] | 3.15E-01 | 0.9 [0.71-1.11] | 2.05E-01 |
| vitamt | Number of multi-vitamins subject takes per week | 0.95 [0.93-0.97] | 3.51E-15 | 0.94 [0.89-1.01] | 8.97E-03 | 0.87 [0.79-0.98] | 4.87E-04 |
| vitb6 | Subject regularly takes Vitamin B6 | 1.0 [0.97-1.03] | 9.94E-01 | 1.04 [0.93-1.16] | 4.04E-01 | 0.95 [0.76-1.16] | 5.12E-01 |
| vitbcom | Subject regularly takes B-Complex vitamin | 0.96 [0.94-0.99] | 4.07E-04 | 0.92 [0.84-1.02] | 2.63E-02 | 0.88 [0.72-1.04] | 5.10E-02 |
| vitbetac | Subject regularly takes beta-carotene | 0.96 [0.89-1.05] | 2.12E-01 | 1.07 [0.8-1.34] | 4.94E-01 | 0.88 [0.47-1.32] | 4.54E-01 |
| vitc | Subject regularly takes Vitamin C | 0.96 [0.94-0.99] | 2.74E-05 | 1.04 [0.96-1.13] | 1.71E-01 | 0.96 [0.81-1.11] | 4.00E-01 |
| vitca | Subject regularly takes Calcium | 1.15 [1.13-1.18] | 2.36E-56 | 2.11 [1.97-2.27] | 2.21E-177 | 1.89 [1.67-2.16] | 2.39E-43 |
| vitcod | Subject regularly takes cod liver oil | 1.0 [0.95-1.05] | 9.31E-01 | 0.9 [0.76-1.06] | 8.12E-02 | 0.91 [0.65-1.2] | 3.63E-01 |
| vitcop | Subject regularly takes copper | 0.93 [0.85-1.02] | 3.11E-02 | 0.96 [0.7-1.25] | 7.00E-01 | 0.78 [0.4-1.23] | 2.02E-01 |
| vitd | Subject regularly takes Vitamin D | 1.03 [1.01-1.05] | 2.37E-05 | 1.27 [1.19-1.36] | 3.90E-22 | 1.11 [0.98-1.26] | 2.27E-02 |
| vite | Subject regularly takes Vitamin E | 0.95 [0.92-0.98] | 1.44E-06 | 0.93 [0.84-1.02] | 5.84E-02 | 0.91 [0.75-1.09] | 1.40E-01 |
| vitfa | Subject regularly takes folic acid | 0.92 [0.89-0.96] | 2.35E-08 | 0.98 [0.86-1.12] | 6.72E-01 | 0.89 [0.7-1.11] | 1.88E-01 |
| vitiod | Subject regularly takes iodine | 0.97 [0.89-1.05] | 3.49E-01 | 1.19 [0.89-1.52] | 7.12E-02 | 0.96 [0.5-1.49] | 8.17E-01 |
| vitiron | Subject regularly takes Iron | 1.0 [0.97-1.03] | 8.77E-01 | 1.03 [0.92-1.15] | 3.99E-01 | 1.06 [0.86-1.27] | 4.22E-01 |
| vitmag | Subject regularly takes magnesium | 0.93 [0.9-0.97] | 2.21E-08 | 1.03 [0.91-1.15] | 5.55E-01 | 0.97 [0.77-1.19] | 6.85E-01 |
| vitmetcl | Subject regularly takes Metamucil | 1.0 [0.95-1.04] | 7.80E-01 | 0.83 [0.69-0.97] | 2.23E-03 | 0.79 [0.58-1.0] | 1.98E-02 |
| vitmv | Subject currently takes multi-vitamins | 0.98 [0.96-1.01] | 3.86E-02 | 0.87 [0.81-0.93] | 7.47E-08 | 0.81 [0.71-0.93] | 1.05E-05 |
| vitnia | Subject regularly takes niacin | 0.95 [0.9-0.99] | 8.58E-04 | 0.93 [0.79-1.08] | 1.79E-01 | 0.95 [0.7-1.23] | 6.25E-01 |
| vitomg3 | Subject regularly takes Omega-3 fatty acid | 0.98 [0.95-1.0] | 1.42E-02 | 0.93 [0.85-1.01] | 3.00E-02 | 0.82 [0.69-0.96] | 1.11E-03 |
| vitoth | Subject regularly takes other vitamins or supplements | 0.98 [0.95-1.01] | 7.92E-02 | 0.89 [0.8-0.97] | 9.33E-04 | 1.12 [0.95-1.31] | 5.62E-02 |
| vitsel | Subject regularly takes Selenium | 1.03 [0.97-1.09] | 1.87E-01 | 1.29 [1.1-1.53] | 2.32E-05 | 1.01 [0.7-1.34] | 9.48E-01 |
| vityrs | Number of years subject has taken multi-vitamins | 0.95 [0.94-0.97] | 1.80E-22 | 0.84 [0.81-0.88] | 1.61E-24 | 0.83 [0.77-0.9] | 3.70E-10 |
| vitzinc | Subject regularly takes Zinc | 0.93 [0.9-0.97] | 7.25E-06 | 0.97 [0.84-1.1] | 5.26E-01 | 0.96 [0.75-1.21] | 6.31E-01 |

**Supplementary Table 4: Exploratory univariable analysis of 126 baseline diet questions and association with three PCa clinical endpoints.** Cox proportional hazards model results from association with age at prostate cancer, metastatic prostate cancer (nodal or distant) and death from prostate cancer. Hazard ratios (HR) are calculated per unit. Answers for average consumption in the last year were on a 9-point scale (Never/less than once a month, 1-3 times per month, once a week, 2-4 per week, 5-6 per week, once a day, 2-3 per day, 4-5 per day 6+ per day). *P*-values reported are from univariable models for each variable. No correction for multiple comparisons was applied for this exploratory analysis. Results should be interpreted cautiously.
